# Diagnosis of protozoa diarrhoea in *Campylobacter* patients increases markedly with molecular techniques

**DOI:** 10.1101/2023.01.05.23284190

**Authors:** David T S Hayman, Juan Carlos Garcia-Ramirez, Anthony Pita, Niluka Velathanthiri, Matthew A Knox, Paul Ogbuigwe, Michael G Baker, Kamran Rostami, Jan Deroles-Main, Brent J. Gilpin

**Affiliations:** Massey University, Palmerston North, New Zealand; Waikato District Health Board, Hamilton, New Zealand; University of Otago, Wellington, New Zealand; MidCentral District Health Board, Palmerston North, New Zealand; Medlab Central Ltd, Palmerston North, New Zealand; Institute of Environmental Science and Research Ltd. (ESR), Christchurch, New Zealand

## Abstract

**Background:** *Cryptosporidium* and *Giardia* are major food and water-borne causes of diarrhoea globally, and two of the most notified infectious diseases in New Zealand. Diagnosis requires laboratory confirmation carried out mostly via antigen or microscopy-based techniques. However, these methods are increasingly being superseded by molecular techniques for diagnostics. Here we investigate the level of protozoa coinfection identified by molecular methods in *Campylobacter* positive samples missed through use of antigen-based assays and then investigated different molecular testing protocols.

**Methods:** We report the findings of two observational studies; the first among 111 people with diarrhoea during a large *Campylobacter* outbreak in Havelock North, and the second a study during normal surveillance activities among 158 people presenting with diarrhoea and a positive *Campylobacter* test, but negative *Cryptosporidium* and/or *Giardia* antigen-based diagnostic test result. The molecular methods used for comparison with the antigen-based tests were in-house end-point PCR tests targeting the *gp60* gene for *Cryptosporidium* and *gdh* gene for *Giardia*. DNA extraction was performed with and without bead-beating and comparisons with commercial real-time quantitative (qPCR) were made using clinical samples diluted down to 10^−5^ for *Cryptosporidium* positive samples.

**Results:** The coinfection prevalence was 9% (n= 10, 3–15% 95%CI) for *Cryptosporidium* and 21% (n=23, 12– 29% 95%CI) for *Giardia* in the 111 *Campylobacter* patients of the Havelock North outbreak. The coinfection prevalence was 40% (n=62, 32-48% 95%CI) for *Cryptosporidium* and 1.3% (n=2, 0.2-4.5% 95%CI) for *Giardia* in the 158 routine surveillance samples. Sequencing identified *Cryptosporidium hominis, C. parvum*, and *Giardia intestinalis* assemblages A and B among patients. We found no statistical difference in positive test results between samples using end-point PCR with or without bead-beating prior to DNA extraction, or between the in-house end-point PCR and qPCR. The qPCR Ct value was 36 (35-37 95%CI) for 1 oocyst, suggesting a high limit of detection.

**Discussion:** In surveillance and outbreak situations we found diagnostic serology testing substantially underdiagnoses *Cryptosporidium* and *Giardia* coinfections in *Campylobacter* patients. These findings suggest that the impact of protozoa infections may be underestimated, through underdiagnosis, but molecular techniques likely improve detection capabilities. Laboratories need to understand clinical, rather than analytical, test sensitivity, to allow clinicians to better understand the disease aetiologies of patients that enable better health advice.

## Introduction

Millions of cases of diarrhoea each year are caused by protozoan parasites *Cryptosporidium* and *Giardia* (1-5). Cryptosporidiosis is often associated with diarrhoea, nausea, vomiting, abdominal cramps, low-grade fever, headache, and fatigue, and is a leading cause of infant mortality in Low to Middle-Income Countries (LMIC) (2, 3, 6, 7). *Cryptosporidium* infections in LMICs increase with poor hygiene and sanitation (8). In wealthy nations, risk factors include contact with diarrheal person/animals, as well as infected food and water (5, 9, 10). *Giardia* is a leading but neglected cause of infectious gastroenteritis worldwide. The symptoms of giardiasis can resemble irritable bowel syndrome (IBS) (11). The diarrhoea can be watery and voluminous, usually resolving within 1 to 2 weeks without treatment, but if diagnosed it is treatable and treatment reduces illness and transmission. Food and water-borne infections with these parasites result in missed school and work and impacts people’s well-being and economies (12).

These diseases are two of the four most notified diseases in New Zealand (13), showing a high annual incidence with relatively little change over the period preceding and including our study (2013-2018), which suggests lack of effective disease prevention and control (Fig 1). Diarrhoea case definition in New Zealand follows the WHO of ≥3 abnormally loose stools in 24 hrs (14).

**Figure 1.**
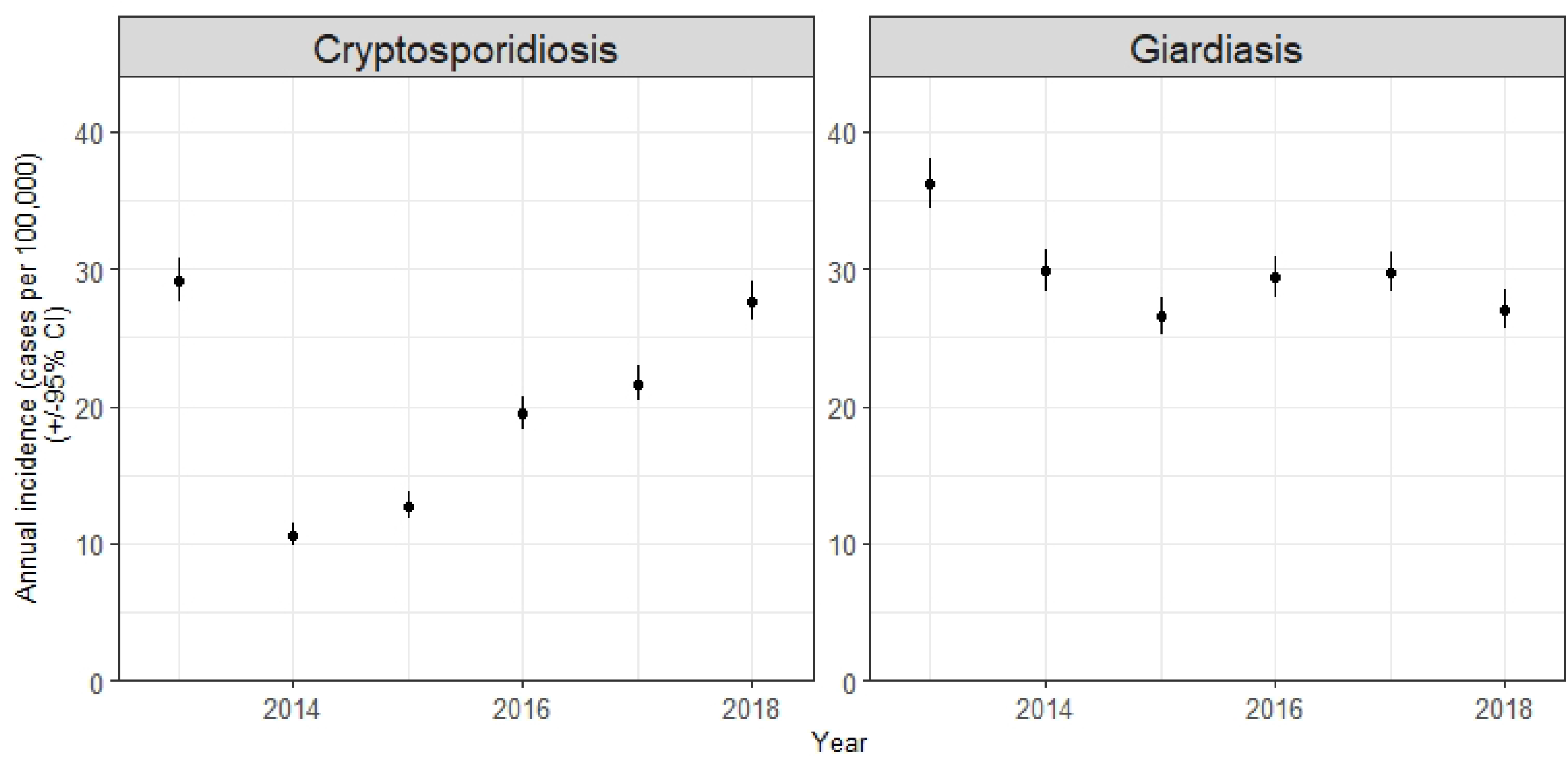
Notified *Cryptosporidium* and *Giardia* prevalence in New Zealand, 2013-2018, covering the study period. Ninety-five percent confidence intervals are shown.

The clinical description of cryptosporidiosis is an acute illness that includes symptoms of diarrhoea (may be profuse and watery) and abdominal pain. The infection may be asymptomatic but to meet the case definition the individual must have compatible symptoms (15). Infection must be confirmed by detection of *Cryptosporidium* oocysts in a faecal specimen using at least one of the following methods: *Cryptosporidium* antigen detection based on direct fluorescence using monoclonal antibodies, a rapid antigen test, or enzyme immunoassay, or the detection of *Cryptosporidium* nucleic acid or visualisation by direct microscopy detection of *Cryptosporidium* oocysts (15).

The clinical description of giardiasis is an illness characterised by diarrhoea, abdominal cramps, bloating, flatulence, nausea, weight loss and malabsorption. The infection may be asymptomatic. Given the remitting/relapsing and variable nature of symptoms, the individual does not need to have compatible symptoms at the time of presentation but must have had a clinically consistent illness to meet the case definition (15). Giardiasis in New Zealand is confirmed by the same methods as *Cryptosporidium*, but for *Giardia* antigen/DNA, and visualisation of *Giardia* cysts or trophozoites (16).

For both diseases, cases are classified as: Under investigation (a case that has been notified, but the information is not yet available to be classified it as probable or confirmed); Probable (a clinically compatible illness that either is a contact of a confirmed case of the same disease or has had contact with the same common source – that is, is part of a common-source outbreak); Confirmed (a clinically compatible illness accompanied by laboratory definitive evidence) or Not a case (a case that has been investigated and subsequently found not to meet the case definition) (15, 16).

Our Protozoa Research Unit (PRU; http://protozoa.org/) at the Molecular Epidemiology & Public Health team at Massey University performs surveillance work on human faeces and water samples for the New Zealand Ministry of Health (17, 18). Molecular genotyping is performed on human faecal samples that have already tested positive through International Accreditation New Zealand (IANZ) diagnostic laboratories (see methods). This work assists public health officers with investigations of outbreaks caused by these and other notifiable diseases, such as a massive outbreak of *Campylobacter* (6260-8320 cases) caused by contaminated water in Havelock North, New Zealand in August 2016 (19).

The Havelock North outbreak provided laboratory evidence of false-negative diagnostic tests for protozoa. To determine if coinfection of *Campylobacter* positive samples were missed due to the test methods under normal surveillance conditions, in 2017 we obtained ethical approval to assess samples from accredited laboratory *Campylobacter* positive, protozoa antigen-assay negative faeces (like the Havelock North outbreak). Therefore, we report the findings of two observational studies. The first study was conducted among people with diarrhoea during the large *Campylobacter* outbreak in Havelock North, which found evidence of coinfection with both protozoa using molecular methods, but not antigen-based methods and the second study as part of normal surveillance activities among people presenting with diarrhoea and a positive *Campylobacter* test, but a negative diagnostic test result for *Cryptosporidium* and/or *Giardia*, replicating the Havelock North test scenarios to determine if this was a common problem. We then report a laboratory study of the impact of DNA extraction technique on our in-house end-point PCR tests and compare a commercial real-time PCR with our in-house PCR.

## Methods

### Ethics

The Havelock North outbreak samples were collected and analysed through routine public health investigation activities. The 2006 guidelines from the National Ethics Advisory Committee, Ministry of Health, Wellington, New Zealand, state that these activities do not require ethics committee review. The second study was approved by Massey University Human Ethics Committee number SOA 17/09. All samples were anonymised clinical samples and patient consent not sought.

### Study location

We present two main study locations. The first is the town of Havelock North, New Zealand. In August 2016 Havelock North had an unchlorinated reticulated water system that supplied water for 14,118 residents, and some residents within approximately 50 km of the neighbouring cities of Hastings (population ∼64,000) and Napier (population ∼61,100). All towns together with other small rural communities (totalling another ∼15,000) comprise the Hawke’s Bay District Health Board (HBDHB) catchment area. The second location is the MidCentral District Health Board (MCDHB) region, including Palmerston North (population ∼85,000), and Tairawhiti and Whanganui DHBs, served by the Medlab Central (total population approximately 280,000).

### Laboratory methods Diagnostic tests

Diagnostic services were supplied by service providers to HBDHB during the Havelock North outbreak and Medlab Central to MCDHB for the routine surveillance study. For the laboratory work, all case samples were processed as normal for enteric pathogens (including *Campylobacter, Salmonella*) based on the clinician’s assessment and test requests (19). Medlab Central Ltd is an IANZ accredited laboratory that used an antigen detection method (Immunocard STAT!® Crypto/Giardia Rapid Assay Kit, Meridian) for protozoa at the time of the studies in 2016 and 2017 respectively.

### Molecular tests

We (PRU at Massey University) received anonymized faecal samples from patients. Samples were stored at 4 °C until processing. We used a bead beater at 30 Hz for 5 min for disruption of cysts (*Giardia*) or oocysts (*Cryptosporidium*) prior to DNA extraction using a Quick-DNA Faecal/Soil Microbe Kit (Zymo Research, Irvine, California, United States). PCR amplification was performed using an in-house nested-PCR assay for *Cryptosporidium* gp60 (60 kDa glycoprotein) and *Giardia* GDH (glutamate dehydrogenase) genes (17, 18), which we used for routine genetic surveillance under the Ministry of Health contract, following diagnosis through IANZ accredited laboratories.

We hypothesized that the DNA extraction method, and in particular the bead-beating to extract DNA, and our in-house test nested PCR led to greater sensitivity for the research laboratory PCR test than immunoassays. To test this, we serially diluted known *Cryptosporidium* DNA positive faecal samples ten-fold from neat to 10^−5^, though only 10^−3^ are reported. We took 5 positive samples and 3 replicates for each. We then extracted genomic DNA using a Quick-DNA Faecal/Soil Microbe Kit (Zymo Research, Irvine, California, United States) with and without the use of a bead-beater (Tissue Lyser II, Qiagen) at 30 Hz for 5 min to disrupt the oocysts. The purified DNA was stored at − 20 °C prior to further processing. PCR was with the same gp60 nested PCR.

We further hypothesised that newer commercially available multiplexed real-time PCRs (qPCR) would be more sensitive than antigen-based assays and a subset (n=19) of the above dilutions were sent to Medlab Central for processing using their new diagnostic Tandem-Plex® Faecal Bacteria and Parasites 12-well (Ref 25041) commercial multiplexed (MT) qPCR. This MT-PCR detects *Salmonella* spp., *Shigella* spp., *Campylobacter, E*.*coli* O157, *Clostridium difficile* toxin A, *Clostridium difficile* toxin B, *Yersinia enterocolitica, Yersinia pseudotuberculosis*, Shiga toxin 1, Shiga toxin 2, *Giardia, Cryptosporidium*, and *Entamoeba histolytica*.

### Sequence analyses

Both strands of the PCR amplification products were sequenced using Big Dye Terminator version 3.1 reagents and an ABI 3730XL automated DNA sequencer (Applied Biosystems, Foster City, California, USA). Consensus sequences were assembled from forward and reverse reads and edited manually using Geneious v.10.1.3 (20). The sequences derived were used to identify species and genotypes by aligning to sequence entries in nucleotide databases using the program BLAST (http://www.ncbi.nlm.nih.gov/blast/) and checked by their corresponding genotype (e.g. (21)). All sequences have been previously reported with accession numbers KY123918–KY124121 and MT265681-MT265802 (17, 18).

### Microscopy

Direct microscopy using BX 60 fluorescence microscope (Olympus; Tokyo, Japan) and Immuno-fluorescent antibody staining using EasyStain (BioPoint Pty Ltd; Sydney, Australia) was carried out in our lab for a subset of samples. Microscopy was also used to quantify the *Cryptosporidium* oocyst counts for the qPCR limit of detection study.

### Statistics

Simple binomial confidence intervals were calculated for all prevalence estimates in R (22) and geometric means and their confidence intervals using the DescTools package (23). A linear model was used to calculate the Ct value cut off in the qPCR test. Further data presentation was done using ggplot2 in R (24).

## Results

### Havelock North outbreak

We (PRU) received 111 samples for testing. All our PCR positive and negative controls were positive and negative respectively, indicating the PCR reactions worked. Ten samples tested positive for *Cryptosporidium* and 23 for *Giardia*, leading to an overall prevalence of 9% (3–15% 95%CI) for *Cryptosporidium* and 21% (12–29% 95%CI) for *Giardia*. We extracted DNA from 15 Giardia positive samples for confirmatory genetic sequence analysis (Fig 2A) which indicated the *Giardia* were from assemblages A and B (Fig 2A). These samples were negative by the antigen-based assay used by the IANZ accredited laboratory serving HBDHB. It was not known if these *Cryptosporidium* and *Giardia* results were related to disease in any cases, as all the samples were positive for *Campylobacter* (25).

**Figure 2.**
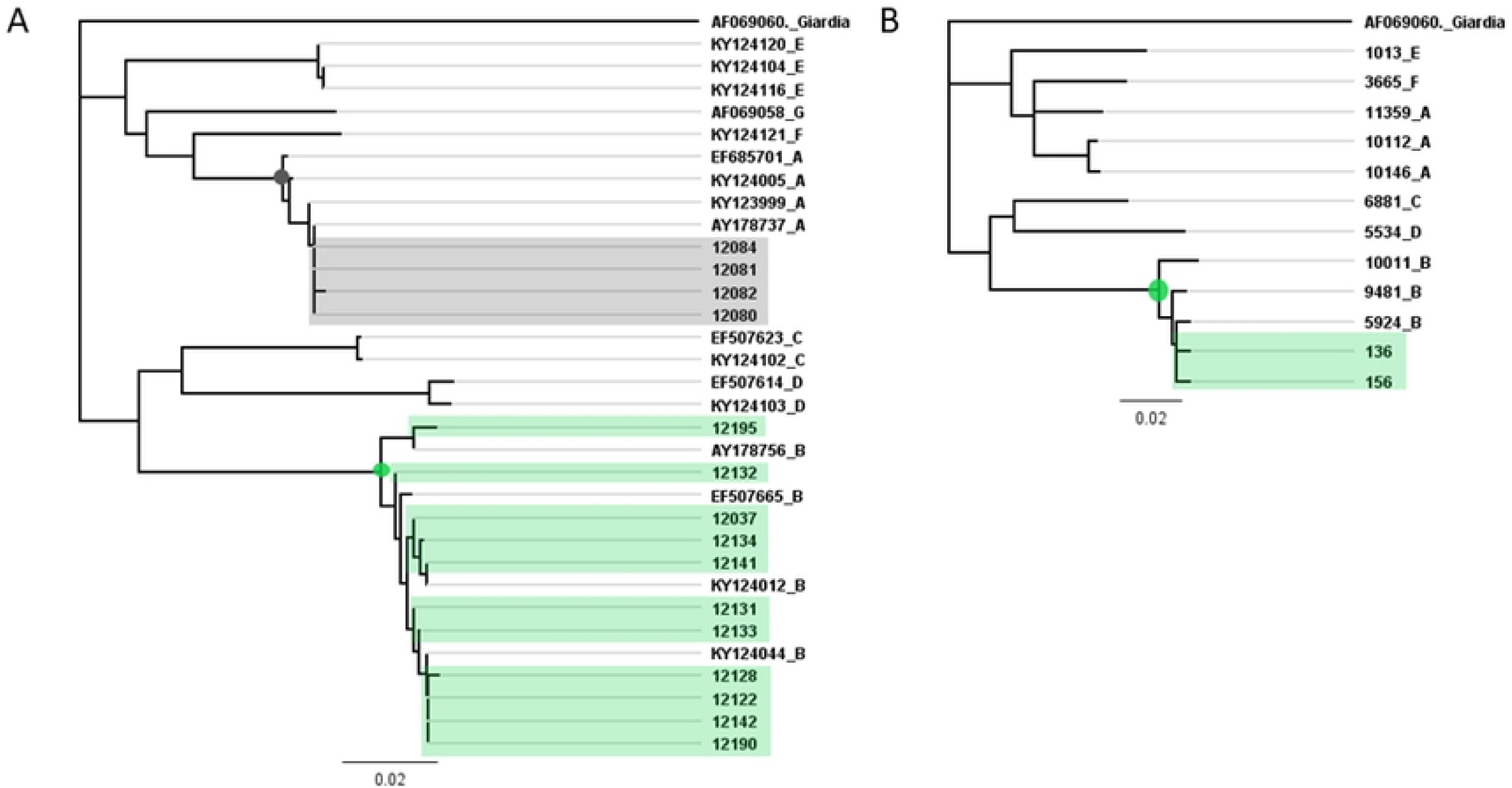
A. Fifteen *Giardia* DNA sequences (GDH gene) from a subset of 23 positive human faecal samples from the Havelock North *Campylobacter* study. *Giardia* from assemblages A (grey node and box) and B (green node and box) were present (25). B. Two of 158 samples from the surveillance study had *Giardia* DNA GDH gene sequences from assemblages A (green node and box). Other reference sequences were obtained from GenBank and accession numbers and assemblages shown.

### Surveillance study

We tested 158 *Campylobacter* positive, protozoa negative faecal samples, to approximately replicate the Havelock North scenario. We detected *Cryptosporidium* DNA from 63 samples (40%, 32-48% 95%CI) and *Giardia* DNA from 2 samples (1.3%, 0.2-4.5% 95%CI). Using sequencing, *Cryptosporidium hominis* was detected in 60 samples (38%, 30–46% 95%CI) with two subtypes (Ib and Ig, Fig 3A) and *C. parvum* with two subtypes (IIa and IId, Fig 3B) (2%, <1%-2% 95%CI). *Giardia* samples all belonged to assemblage B (Fig 2B). Again, all negative controls were negative and positive controls positive.

**Figure 3.**
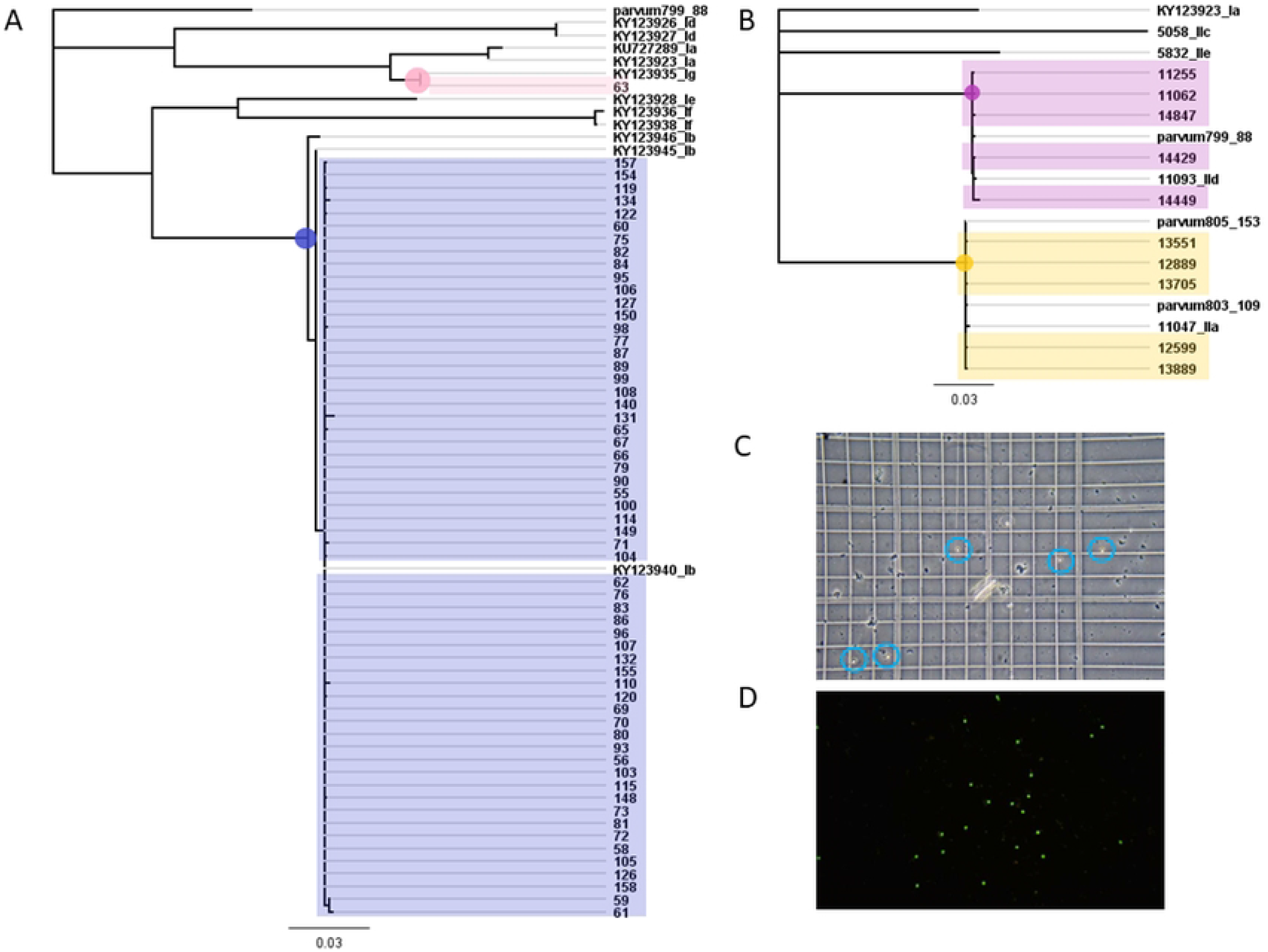
A. Molecular evidence and genetic relationships of the *Cryptosporidium hominis* (n = 60) with two subtypes; Ib (blue node and box) and Ig (pink node and box), and in B. *C. parvum* (n = 3) with 2 subtypes (IIa; yellow node and box, and IId; purple node and box). Other reference sequences were obtained from GenBank and accession numbers and species and genotype (e.g. Ib) are shown.C. Microscopic (see in blue circles) and D. immunofluorescent antibody staining (bright dots) detection of *Cryptosporidium* oocysts in human stool samples.

To further confirm the results, we randomly chose 10 positive samples and confirmed the presence of *Cryptosporidium* oocyst by light microscopy and immuno-fluorescent antibody staining (Fig 3C & 3D).

Overall, from 269 samples that were protozoa antigen-assay negative, *Campylobacter* positive, we detected 86 *Cryptosporidium* (32%, 95%CI = 26–38%) and 12 *Giardia* (4%, 2–8% 95%CI) positive samples. The expected change in notifications nationally if our lab tests were applied to national *Campylobacter* data would make cryptosporidiosis the second most notified infectious disease in New Zealand (Table S1).

### DNA extraction and detection limit comparisons

Contrary to our hypothesis we found no significant difference between our ability to detect *Cryptosporidium* DNA whether we used bead-beating or not (*χ*^2^ = 0.75, p-value = 0.39, Fig 4A) with 36% of the dilution series samples positive (27-46% 95%CI) using bead-beating and 43% (34-53% 95%CI) not. Using a small subset of 19 samples we compared the in-house gp60 gene PCR results to the commercial qPCR and found no significant difference (*χ*^2^=1.2, p-value = 0.27), with 84% (62-94% 95%CI) positive by qPCR and 63% (41-81% 95%CI) by end-point PCR (Fig 4B)

**Figure 4.**
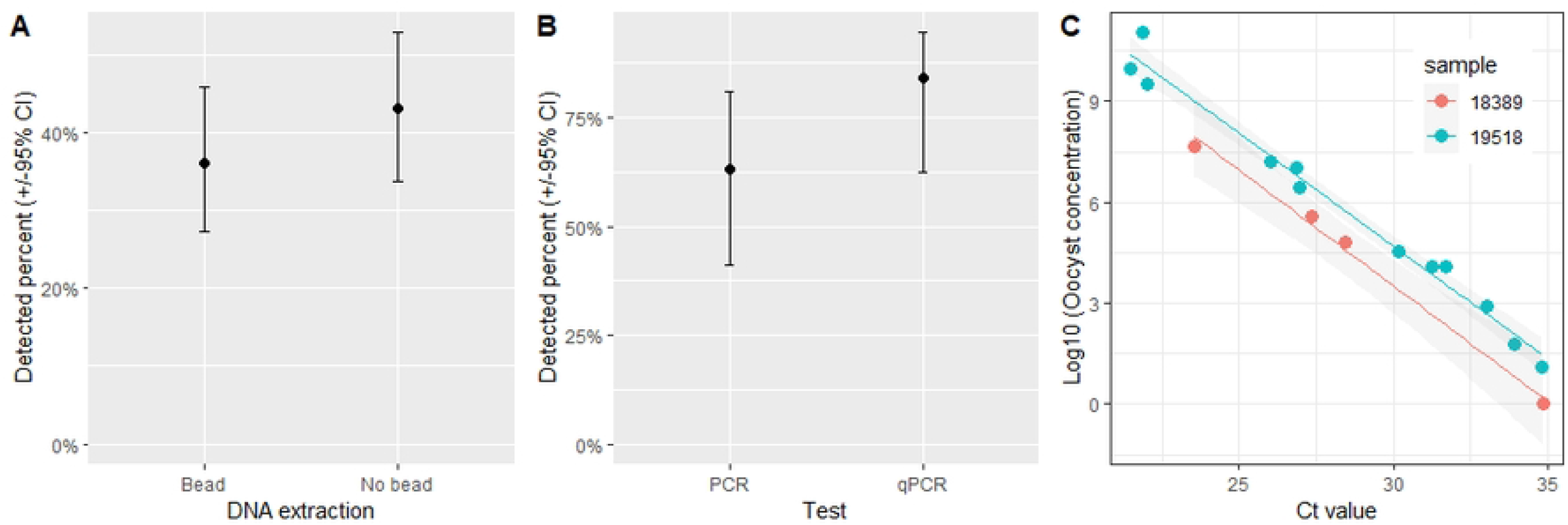
*Cryptosporidium* detection using different molecular methods. A. DNA extracted with (bead, n = 18) and without (no bead, n = 18) bead-beating to damage cells before DNA extraction and an endpoint nested in-house gp60 gene PCR were used. The percent of all *Cryptosporidium* positive faecal samples in ten-fold dilutions to 10^−3^ which detected *Cryptosporidium* is shown with 95% confidence intervals B. Comparison of this in-house end-point gp60 PCR and a commercial qPCR (n=19). C. The correlation between Log10 oocyst concentration and commercial qPCR Ct value from serial dilutions of two positive samples.

The detection limits by endpoint PCR were never greater than 10^−3^ dilutions of faeces in our test. The qPCR Ct values scaled linearly with the log oocyst concentration (Fig 4C), with an estimated Ct value for 1 oocyst of 36.9 (35.9-37.8 95%CI) for one sample (19518), 35.0 (33.0-37.1 95%CI) for another (18389) and 36.2 (35.1-37.2 95%CI) for both.

## Discussion

We identified a large number of protozoa positive samples among 269 *Campylobacter* positive patient faecal samples that tested negative in diagnostic laboratories using antigen-based assays. The prevalence ranged from 1 to 40%, depending on the scenario and pathogen. Our findings indicate that background *Giardia* and *Cryptosporidium* infection incidence is higher than detected and reported, and coinfections are common. Accredited antigen-based tests appeared to be less sensitive than the molecular methods used in the research laboratory during outbreak and surveillance scenarios. The impact of these false negatives in New Zealand is unknown. However, misdiagnosis among diagnostic laboratories globally is likely to be similar and, therefore, the impact and burden of both protozoa potentially greatly underestimated. These findings are likely to have global relevance for both the clinical management of cases which are underdiagnosed, and for public health management of these diseases which is systematically under-estimating their impact.

We (Massey) still use this endpoint PCR for sequencing the gp60 for *Cryptosporidium* and gdh gene for *Giardia* for characterisation, but this is not a diagnostic PCR and is more labour intensive. Other studies have demonstrated microscopy to be less reliable than PCR with 83.7% sensitivity and 98.9% specificity compared to PCR (26) and diagnostic sensitivities of three *Cryptosporidium* and *Giardia* combination enzyme immunoassays (EIA) to be 91.4–93.4 %, with the sensitivity of auramine phenol microscopy as 92.1 % and immunofluorescence microscopy (IFM) 97.4 %, but with no significant difference between them (27). However, we showed in our studies that antigen-based tests were missing samples with levels of oocyst presence easily detectable by microscopy (Fig 3C &3D).

Our hypothesis that bead-beating impacted the test sensitivity and limit of detection was not supported. We perform bead-beating to break the (oo)cyst walls, however different *Cryptosporidium* and *Giardia* life stages are less resilient and may be present in faeces in addition to (oo)cysts and/or vortexing maybe be sufficient to expose the DNA, explaining this finding. Diagnostic laboratories in New Zealand have switched to multiplexed qPCR since our study, which meant we were not able to directly test our results using the dilution series. However, encouragingly, the multiplexed qPCR used by Medlab Central compared well to our endpoint PCR. We showed that the qPCR Ct value correlates well with the log10 concentration of oocysts and can detect as few as 1 oocyst in a diluted sample.

All three pathogens, *Campylobacter* and the two protozoa, can have ruminant hosts (25, 28). In our two ecological studies there were different findings; first in the Havelock North outbreak we found more *Giardia* than *Cryptosporidium*, whereas in the surveillance study this pattern was reversed. This finding may reflect different prevalence of the infections in different populations or that the outbreak was associated with a contaminated water point source which might have also been contaminated with *Giardia* from the same source, compared to the surveillance that was more a reflection of the normal background infection. Given the highly seasonal nature of both infections either scenario may be possible.

In our study all the *Campylobacter* patients had diarrhoea. In the UK, for each *Giardia* case reported to national surveillance there are 14 cases in the community (29). In New Zealand, notified cases are likely extremely low compared to the burden of disease. The 2006-2007 New Zealand Acute Gastrointestinal Illness (AGI) Community Study (12), a representative cross-sectional telephone survey of 3655 participants, reported an incidence of 1·11 AGI episodes per person per year. Prevalence was highest in children aged <5 years, with the most common symptoms being diarrhoea (83%) of 2.5 days. Adjusted estimates mean there were about 4·66 million AGI episodes per year in New Zealand, with nearly 1 million visits to the general medical practitioner with “in excess of 300 000 courses of antibiotics being dispensed and more than 4·5 million days of paid work lost due to AGI” (12). This represents a significant burden of AGI disease for New Zealand and our analyses suggest the role of protozoa may have been underestimated.

Both children and adults with cryptosporidiosis reportedly often have diarrhoea in wealthy countries (5, 6). However, in LMIC diarrhoea or other symptoms are only reported in ∼1/3 of *Cryptosporidium*-infected children (30), though subclinical cryptosporidiosis can have significant adverse effects on children (31-34). Asymptomatic carriage of *Giardia* is common in contacts of cases. In both *Cryptosporidium* and *Giardia*, however, there is a correlation between the type of infection (species, assemblage, genotype) and the presence of symptoms (35). For example, in Saudi Arabia children infected with *G. intestinalis* B assemblages were symptomatic, while asymptomatic children had only assemblage A (36). We discovered both in patients. Similarly, co-infection, including with other *Cryptosporidium*, was more likely to lead to disease (2, 3). We have demonstrated that coinfection with *Campylobacter* was common in our Havelock North and surveillance data. In recent work, we have developed a “metabarcoding” method and discovered coinfection by multiple *Cryptosporidium* species and genotypes (unpublished) and *Giardia* assemblages (37). This observation poses the question; does carriage lead to an increased risk of disease when a secondary infection or coinfection occurs?

It is not clear if treatment of asymptomatic protozoa carriage is effective in limiting transmission (38), but findings have raised the public health question of whether asymptomatic household contacts should be treated to prevent infection (29). *Giardia* infections have been discovered in 19-30% of households with cases with the presence of gastrointestinal illness in the household before the onset of the index case a risk (e.g. odds ratio 9; 95% confidence intervals 1.5–48) (36). We also expect contact with *Cryptosporidium*-positive patients to lead to secondary transmission in households (39, 40). Case-control studies of *Cryptosporidium*-infected people in Asia and Africa have found the secondary infection rates from 8 to 36% (40, 41), with the differences perhaps attributable to the dominant *Cryptosporidium* species or subtype present. Furthermore, at least in LMICs, children can experience multiple episodes of cryptosporidiosis (5, 42), with some reports of up to four episodes before the age of two years, with no decreases in parasite burden during repeat infections (30). This evidence suggests that even if not related to clinical disease, missing protozoa on tests may contribute to ongoing transmission and health issues.

There are key questions in *Giardia* research that are also relevant to our findings (43): Do different *Giardia intestinalis* genetic assemblages differ in transmission and clinical outcome of disease? Is previous exposure to *Giardia* protective against re-infection? What is the relationship between *Giardia* and IBS or other post-infectious gastrointestinal disorders? Is the treatment of asymptomatic cases necessary to reduce transmission? Similar questions exist for *Cryptosporidium*. It would also be useful to repeat this testing using diarrheal specimens where pathogens other than *Campylobacter* were detected, and also where no pathogens were found, to improve our estimate of the contribution of these protozoan pathogens to enteric disease more generally. False-negative results will limit the ability of researchers and clinicians to address these questions. More sensitive molecular methods with parasitology panels are increasingly used by diagnostic laboratories, with some evidence of improved detection (44, 45). Thus, our work, suggests improving laboratory methods and case definitions may enhance case diagnosis, resulting in better clinical and public health management of these diseases.

Molecular methods are still not universally available in New Zealand, nor are protozoa tests always requested by clinicians. But these will be even less available in resource poor countries. Our results will help laboratories understand the test’s clinical sensitivity (i.e., case detection), rather than analytical sensitivity (i.e., dilution measurement). Our results suggest clinicians must determine the extent to which tests results can be relied on. Diagnosis might mean more than one test is required if clinical symptoms persist and test results are negative, including as diagnostic laboratories make the transition to molecular testing (43-45). For example, Medlab Central has just converted to PCR and the impact of the transition is to be seen. However, the strict Covid-19 restrictions has reduced the circulation of *C. hominis* in New Zealand, whereas *C. parvum* has continued to cause disease (28). Therefore, it will be some time before the impact of changes to the laboratory testing can be seen in New Zealand.

Asymptomatic *Giardia* infection is generally not treated, but treatment may be rational if asymptomatic cases lead to onward transmission. The wider availability of sensitive PCR diagnostic tests may allow a more targeted approach to contact treatment in future. In secondary care when giardiasis is highly suspected but stool results are negative, diagnosis can be made through duodenal aspiration and biopsy, which have been shown to detect infection in the absence of cysts on stool microscopy (43, 46, 47); improvements in test sensitivity may reduce this need. The symptoms of giardiasis can resemble IBS (11), a condition affecting 1 in 7 people in New Zealand, and IBS or dyspepsia patients may have *Giardia* (e.g. 6.5% of patients (48), but see (46)). The UK National Institute for Health and Care Excellence recommends that faecal testing for parasites is not routine when confirming IBS in people who meet the diagnostic criteria (43), but that clinicians should be aware of both diagnoses. Improved diagnostics will help clinicians in New Zealand determine when a parasitological examination of a stool sample is required (43).

Advice on protozoa for patients and carers will be incorrect with false-negative tests, so our work will help the Ministry of Health, clinicians and community workers reinforce simple health advice, such as handwashing, along with other measures, e.g. for those with *Cryptosporidium*, for which there is no treatment, not use public swimming pools until 2 weeks after symptoms have resolved, rather than the standard 48 hr AGI recommendation, nor should they work with immunocompromised people during this period, including elderly and young (43).

Overall, our results suggest that protozoan diarrhoea is likely significantly underdiagnosed. The switch to sensitive qPCR in diagnostic laboratories is likely to improve diagnosis and help reduce false negatives. However, there is still work to be done to improve test sensitivity and understand the impacts of false negative results on transmission and disease.

## Data Availability

Data are available in the manuscript or publicly available via GenBank and accession numbers are provided.

## Acknowledgements

The authors would like to thank Drs Chris Hewison, Nicholas Jones, and Tom Kiedrzynski for their work on the Havelock North outbreak and Tom for comments on this work.

## Supporting information

Table S1: The actual (left) and adjusted (right) case notifications of the most notified infectious diseases in New Zealand in 2016. Adjusted data are based only on the differences between our research laboratory and the accredited diagnostic laboratories diagnostic results for *Campylobacter* positive but protozoa negative test results.

